# Managing Knowledge in Computational Models for Global Food, Nutrition and Health Technologies

**DOI:** 10.1101/2020.06.05.20122952

**Authors:** Kudakwashe Dube, Scott McLachlan, Ngonidzashe Zanamwe, Evangelia Kyrimi, Jasmine S Thomson, Norman Fenton

## Abstract

Computational models that need to incorporate domain knowledge for realistic solutions to problems often lead to technologies that get transferred to developing countries. The support for managing the knowledge incorporated into these technologies is important for customisation to suit local conditions. This work investigates this problem through the challenge of incorporating *food and nutrition therapy guidelines* (FNTG) into ICT-based solution models for the *meal planning problem* (MPP) for HIV/AIDS patients in developing countries. An experiment is undertaken to demonstrate the limitations of existing approaches and framework is investigated for manageable incorporation of knowledge into solution models. The paper contributes a clear understanding of, and directions for addressing, the problem of support for managing the knowledge incorporated into solutions models to support customisability of technologies. The significance of this contribution is that solutions will allow resulting technologies to be customised for use in different global contexts.

## I INTRODUCTION

Ensuring that the domain knowledge being incorporated into computational models is manageable is a challenge which requires supporting customisation of the knowledge. This would allow any resulting technology to be customised and used in different global contexts. All computational solution models require domain knowledge to produce *realistic* solutions [1, 2]. Simple problems are solved using little domain knowledge while hard problems require more comprehensive domain knowledge [3, 4]. For optimisation problem-solving methods, domain knowledge is often incorporated in computational models used in their heuristics for coping with the computational complexity of hard problems [5, 6]. The knowledge must first be formalised into computer interpretable form before it can be incorporated for use within a computational model. This requires consideration of the management in order for the resulting technology to be customisable. To be ***manageable***, one should be able to manipulate the incorporated knowledge through defined and supported operations. This would allow customisation of the resulting technology to suit different global circumstances and enables technology transfer.

In this paper, the research problem under investigation is the *manageable incorporation of domain knowledge into computational solution models*. The objectives of the paper are to show that: (1) existing computational models for solving the ***Meal Planning Problem* (MPP)** produce unrealistic solutions without incorporating food and nutrition knowledge; (2) existing computational models do not have inherent or natural ways for supporting *manageable incorporation* of knowledge; and, (3) knowledge incorporated into computational models using existing methods is generally difficult to manage, making them hard to use in different global contexts.

*To investigate the research problem, the paper uses* the MPP in the context of treating a clinical condition, which requires ***Food and Nutrition Therapy Guidelines* (FNTG)** as the domain knowledge. The FNTG give clinicians, dietitians, nutritionists and caregiver’s concise evidence-based instructions on optimal *food and nutrition therapy administration* (FNTA). FNTGs improve the quality and consistency of FNTA because they offer recommendations on how to proceed according to best practice and alert domain experts when a wrong practice has been followed [7]. Formalisation of FNTGs is a challenging problem that requires at least two areas of expertise – the domain expert: a nutritionist, and a knowledge engineer. The domain expert must correctly interpret the FNTG for the knowledge engineer who then formalises the knowledge into computer interpretable format [7].

As part of attaining its objectives, this paper investigates a framework for manageable incorporation of knowledge into computational models. The paper also presents a new understanding on the management challenges for knowledge incorporation into computational models. The framework and management challenges are evaluated through its application to assess existing literature and through an experiment applying the **genetic algorithm (GA)** to the MPP. The paper is organised as follows: first, the relevant literature is reviewed; second, the MPP and food and nutrition therapy guidelines are defined. After that, the theoretical contributions are presented before the design of experiments is outlined. This is followed by presentation and examination of the results and discussion of the significance, implications, and future work are discussed before the paper is concluded.

## II THE CHALLENGE OF SUPPORTING FOOD AND NUTRITION THERAPY FOR HIV/AIDS PATIENTS IN SELF-MANAGED AND HOME-BASED CARE IN DEVELOPING COUNTRIES

HIV/AIDS and poor nutrition are inextricably linked in a vicious cycle. Poor nutrition and malnourishment increases the risk of transmission, and in turn the HIV infection attacks the immune system and interferes with nutrient intake, absorption and utilisation. [8,9] In this way HIV exacerbates malnutrition. Global research has shown that evidence-based nutritional interventions that aim to increase energy and protein intake for the HIV patient, may help to mitigate their vulnerability to weight loss and wasting away. [8,9]

Food and nutrition therapy administration (FNTA) is important and integral in the management of HIV/AIDS patients in both self and managed care [10, 11], especially in Developing Countries where weight loss is widespread due to malnutrition [12]. According to Koethe et al [13] and Martinez et al [14], in Sub-Saharan Africa and Honduras, there are high death rates and low health outcomes in HIV/AIDS patients who start anti-retroviral therapy (ART) with low body mass index (BMI), which mandates FNTA under severe personal and organisational resource limitations. Further to this, Cark and Cress [15] point out that FNTA should be personalised in order to be effective for HIV/AIDS patients, which requires any appropriate technologies to support customisation of incorporated knowledge. The major challenge faced by HIV/AIDS patients in developing regions is the combination of the disease, poverty, hunger and the resource limited environment [16]. Furthermore, lack of food and nutrition is strongly correlated to mortality and adherence to treatment therapies [14].

It is well known that a large number of clinical tests, signs and symptoms must be assessed in order to diagnose that a HIV/AIDS patient has a nutritional deficit requiring therapy and then to devise the most appropriate treatment strategy for supporting that patient’s needs. [17] There is a recognised need for *decision support tools* that can support the diagnosis, monitoring and treatment strategies. [17, 18] *In developing countries*, mobile ICT-based FNTA tools are important in alleviating the challenges in the tasks of therapy personalisation, managing and monitoring patients under the therapy programmes in resource limited settings.

This paper is part of ongoing work to create a technological foundation for mobile ICT-based tools for HIV/AIDS Nutrition Therapy in developing countries with a focus on facilitating transfer of technologies with knowledge components that can be customised to suit both the local environment and the patient. This is expected to result in people living with HIV/AIDS easily, quickly and cost effectively accessing information about food and nutrition therapy programmes through mobile devices thereby improving their health.

## III THE MEAL PLANNING PROBLEM AND FOOD AND NUTRITION THERAPY GUIDELINES

The MPP is a multi-objective and multi-constrained optimisation problem that is often solved by using computational models from the domain of *evolutionary computation* such as the GA. Solving the MPP involves designing a set of meals from food ingredients of different nutritional values. The MPP is traditionally recognised as an intractable problem and guideline knowledge is difficult to formalise and incorporate [19]. The classic MPP is modelled as the *cost minimisation diet problem* [20], often defined mathematically as a *Linear Programming* (LP) problem [21] and a multi-objective optimisation problem [22]. Therapeutic meal planning needs to consider national and international FNTG. Most computational solution models for the MPP generally do not inherently support incorporation and manipulation of knowledge in the form of FNTG.

*Nutrient recommendations* are produced by nutrition experts as sets of evidence-based standards that define the energy and nutrient recommendations for health. Table I shows an extract from a nutrient recommendation [23] used to plan and assess the nutrient intakes of healthy people. Recommendations differ by populations, so they are classified into groupings by gender, age group, and life-stage (for example, pregnancy, lactation).

*Food and Nutrition Therapy Guidelines* (FNTG) generally consist of a list of goals to address diet related health problems and are often expressed as statements promoting change from the current average national diet, for example; *“choose and/or prepare foods that are low in salt”*. FTNG often include physical activity and exercise recommendations. FNTGs are also a type of *Clinical Practice Guidelines* (CPGs), which are important tools for improving outcomes in preventative, curative and, therapeutic care [23, 24].

The work in [7] describes *active* and *passive* approaches to *dissemination* of CPGs. The passive approach involves publishing guidelines in journals. Weaknesses of this approach are: (1) Practitioners might not be aware of the existence of the guideline; (2) Those that are might not be able to locate or use them; and (3) The complexity of some FNTGs render their use very difficult in the clinical setting. For example; the guideline from which Table 1 was extracted from a larger version that spans more than 400 pages. The active and effective approach incorporates guideline knowledge into *decision support systems* (DSS) [7]. Formalisation of FNTGs into computer interpretable format makes this possible [25].

**Table 1:** EXTRACT OF NUTRIENT RECOMMENDATIONS (NATIONAL ACADEMY OF SCIENCES)

| Life Stage Group | Total Water<sup>a</sup> (L/d) | Carbo-hydrate (g/d) | Total Fiber (g/d) | Fat (g/d) | Linoleic Acid (g/d) | α-Linolenic Acid (g/d) | Protein<sup>b</sup> (g/d) |
| --- | --- | --- | --- | --- | --- | --- | --- |
| **Infants** |  |  |  |  |  |  |  |
| 0–6 mo | 0.7* | 60* | ND | 31* | 4.4* | 0.5* | 9.1* |
| 7–12 mo | 0.8* | 95* | ND | 30* | 4.6* | 0.5* | 11+ |
| **Children** |  |  |  |  |  |  |  |
| 1–3 y | 1.3* | 130 | 19* | NDc | 7* | 0.7* | 13 |
| 4–8 y | 1.7* | 130 | 25* | ND | 10* | 0.9* | 19 |

## IV RELATED WORK

Few works investigate manageable incorporation of knowledge in computational models. In the area of the computerization of **clinical practice guidelines (CPG)**, a form of evidence-based knowledge incorporated into clinical care process technologies, one of the authors of this paper introduced the *management/manipulation plane* into the SpEM Framework for managing computerized CPG [25, 26]. Webber and Wu [27] presents a framework for knowledge management defining four knowledge operations: *create, understand, distribute* and *reuse*, which operate at process level and require further validation in practice. The operations do not manipulate *granular* elements of the incorporated knowledge. Furthermore, the knowledge being incorporated is not highly targeted, that is, specialised knowledge about specific diseases or patients.

Managing knowledge incorporated in evolutionary computational models has been recognized as a challenge worthy of investigation. [28] presents an approach supporting only two operations: *share* and c*reate*. In [29], there is a proposal for a *cultural algorithm* in which members of the population acquired, encoded and stored knowledge in a way which allowed knowledge sharing by all members of a population. In [30] there is proposal for a Case-Initialized GA for knowledge extraction and incorporation in which new knowledge could be *created, retrieved* and *updated*. Also, an approach to knowledge incorporation into GAs which only allows for creation of knowledge and application of the knowledge-based mutation operator was proposed in [31]. Generally, in GA, knowledge is incorporated in the fitness function, initialisation process and genetic operators [32]. [33] incorporates knowledge in *representation, population initialisation, recombination* and *mutation, selection* and *reproduction* and *fitness evaluations*. Comparable methods are found in [34–36], with the latter standing out in that it has *knowledge-based initialisation, crossover mutation* and *selection* as methods of knowledge incorporation. All these approaches propose schemes for managing incorporated knowledge that are tightly integrated with the model and hence not sufficient for the problem of this paper. They also do not assist in creating solutions that meet the definition of management of the incorporated knowledge as proposed in our previous paper [37].

Several studies have investigated solutions to the MPP. These solutions differ but can be characterised by the framework used in modelling the MPP and the extent to which they incorporate different types of knowledge. Recently, *three ways for the classification of solution models* [7, 38, 39] for the MPP have emerged in literature.

The **first** identifies four classes that are used in automated menu planning approaches: *trial-and-error, optimisation, metaheuristic* and *fuzzy reasoning approaches* [38]. Most works cited in [38] did not focus on manageable incorporation of food and nutrition therapy guideline knowledge into solution models suggesting research opportunities remain in this area.

The **second** relies on four major categories of computational models [7]: *rule-*based*, document-based, decision-logic expression languages*, and *task-network models*. However, none provides an approach for supporting formalisation of complete food and nutrition therapy guidelines.

The **third** and last arose from the work of the authors of this paper [39]. It classifies solutions depending on whether they are based on *nutrition, mathematical, computational* or *hybrid* model [39]. *Nutrition models* are problem-specific and are usually embodied in a nutrition guideline and tend to be mapped onto either the mathematical or computational model, or both. *Mathematical models* are based on some mathematical formulation such as **Linear Programming (LP)** which has been used to model the MPP [40, 41]. *Computational models* use relevant computing paradigms and formalisms such as: GA [22], Quantum Particle Swarm Optimisation [42], Ant Colony Optimisation [42, 43] and Knapsack Problem [44]. *Hybrid models* integrate two or more models [29, 37, 45].

In the literature, these models do not provide support for managing the incorporated knowledge and hence do not allow for customisation of resulting technologies to suit different contexts. This work therefore investigates this gap in literature with the aim of achieving manageable incorporation of knowledge into solution models for the MPP.

## V FRAMEWORK FOR MANAGING KNOWLEDGE INCORPORATED INTO SOLUTION MODELS FOR FOOD AND NUTRITION THERAPY

### A Solution Models for the MPP

The FNTGs are incorporated into solution models for the MPP according to the level of generality or specificity of the knowledge with respect to the target population. Table II shows that knowledge is incorporated into solution models at four levels, namely, L_0_, L_1_, L_2_ and L_3_ [39]. L_3_ incorporates highly-targeted knowledge. This knowledge is very specific to the target population. In practice, L_3_ does not preclude L_2_, which, in turn, does not preclude L_1_. This means a guideline about a specific disease or patient at L_3_ does not preclude the guideline at L_2_ or L_1_. This is so because at L_3_, although a guideline has more detailed and specific information and knowledge that applies to specific circumstances, guidelines that apply in general circumstances are still applicable. Table 2 also identifies the knowledge element classification for MPP solution models. KD0 represents models at knowledge incorporation level L_0_ which do not incorporate any knowledge and hence not useful in food and nutrition therapy. KD1 represent generic models at knowledge incorporation level L_1_. These are not specific to a health problem or population but are useful in nutrition for general health. Each element may also incorporate the knowledge contained in the previous levels. This means that models at level L_3_ can be used in conjunction with knowledge from general models for specific groups of people (L_2_) and for healthy people (L_1_). However, the models at L_0_ could not be used in place of more advanced models since they do not possess any knowledge at all.

**Table II:** KNOWLEDGE INCORPORATION LEVELS

| Level | Definition | Description | Knowledge Element Classification |
| --- | --- | --- | --- |
| L<sub>0</sub> | NK: No knowledge | No knowledge is incorporated | **KD0:** Model not useful in nutrition therapy |
| L<sub>1</sub> | NTK: Non-targeted knowledge | Knowledge not specific to a particular disease/patient is incorporated | **KD1:** General models for healthy people |
| L<sub>2</sub> | TK: Targeted knowledge | General guidelines about a specific disease/patient but are not identifiable and manageable as a complete nutrition therapy guideline. | **KD2:** General models for specific groups of people |
| L<sub>3</sub> | HTK: Highly targeted knowledge | Complete guidelines about a specific disease/patient are incorporated | **KD3:** Specialised models for patients and specific diseases |

### B Manageable Incorporation of Knowledge into Solution Models for the MPP

Manageable incorporation of knowledge into solution models requires allowing flexibility in form of customisation of the knowledge incorporated into that model. This is necessary to support technology transfer and adaptation. A manageable solution would support incorporation of knowledge elements specified in Table II while also supporting the basic and advanced knowledge manipulation operations presented in Table III. On one hand, there are models which are completely unmanageable while, on the other hand, there are models that are completely manageable. In between, there are models with low, medium and high levels of management of incorporated knowledge. Solution models could incorporate knowledge elements from the knowledge dimensions presented in Table II. If a solution model incorporates elements from any one of these knowledge element classifications but does not support any management operations, that solution model is deemed not to support management of incorporated knowledge. Such solution models constrain flexibility in the customisation and so fails to support technology transfer. Hence, such solution models are not very useful in food and nutrition therapy.

**Table III:** KNOWLEDGE MANIPULATION OPERATIONS

| Operation Class | Operation | Description |
| --- | --- | --- |
| Basic knowledge manipulation operations (cf. CRUD) | *Create* | Generate a new knowledge base from guidelines |
|  | *Retrieve/Query* | Answer requests for knowledge |
|  | *Change/update* | Modify knowledge base to accommodate changes in guidelines |
|  | *Delete/Remove* | Delete knowledge from the knowledge base |
| Advanced knowledge manipulation operations | *Customise* | Modify model and knowledge base to suit specific scenario |
|  | *Replace* | Replace guideline with different one |
|  | *Share* | Export generic version of knowledge base |

If a solution model supports all or most of the operators in Table III, then, for the purpose of this paper, it could be said to support complete management of the incorporated knowledge. Thus, the model could be adapted for use in different regions of the world. Such models are more suitable for patients or specific diseases and are ideal for effective dissemination of guideline knowledge in environments where guidelines are constantly changing or varied. *It should be noted that, within the meal planning therapy domain*, there are models that only support basic operations. Such models are suitable only for healthy people since the knowledge incorporated in the model is not comprehensive enough to be highly targeted. There are also models that support either one operation represented using regular expression terminology as ([share, replace, customise]{1}) or two operations ([share, replace, customise]{2}). The larger the number of operations supported by a solution model, the easier it is to manage knowledge, and the more useful the model becomes to patients and specific diseases.

*The ideal situation is to have MPP solution models that support full management of incorporated knowledge that is highly targeted to a specific health condition*. Fig. 1 shows that most works modelling the MPP using GAs except [46] are using approaches to knowledge incorporation which are not manageable because they did not define any knowledge management operations beyond the basic operations. Most of these works either incorporated NK or NTK in the GA. [46] is the only work falling in the class of models that are hard to manage from the knowledge perspective since it has not defined some basic operations. Therefore, it can be concluded that, existing approaches do not support manageable incorporation of knowledge into GAs for the MPP.

**Fig. 1.**
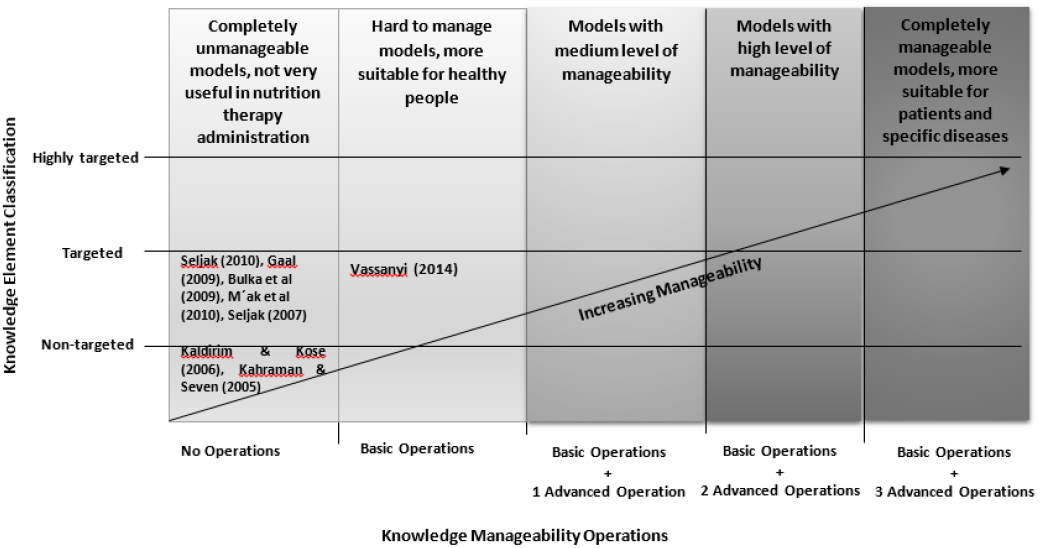
Application of characterisation model for manageable incorporation of knowledge into Genetic models for the MP

## VI EXPERIMENTAL INVESTIGATION OF THE KNOWLEDGE MANAGEMENT CHALLENGE

### A Experiment Approach

An experiment was undertaken to investigate management challenges of guideline knowledge incorporation into the GA-based solution model for the MPP. GA were chosen because most works in literature modelling the MPP used GAs in which some knowledge was incorporated into the GA algorithms [32]. Fig. 2 shows the overall high-level design of experiments. In this design, the user provides personal data as input to the system. Upon receiving the personal data input, the system also receives knowledge from the knowledge base. The system then generates meals and exports them to an external file which is then accessed by the user to get the generated meals. Knowledge was incorporated in GA operators, namely, *crossover, mutation* and *selection*. The experiment modelled the MPP using GA and a Python-based framework called DEAP [47]. The experiment had two parts (P_1_ and P_2_). Crossover is one of the Genetic operators modified to incorporate knowledge.

**Fig. 2.**
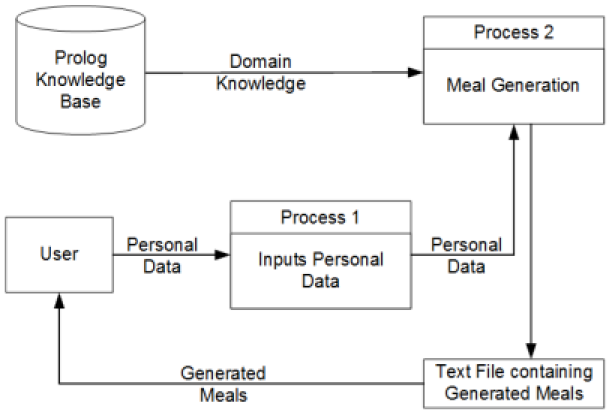
Overall design of Experiments P_1_ and P_2_

Experiment P_1_ sought to show that GAs do not solve the MPP in their natural form without incorporating much guideline knowledge. This was demonstrated by implementing the GA without incorporating much domain knowledge in the Genetic operators of the algorithm. In P_1_, personal data and Food Composition Data (FCD) were incorporated in some genetic operations while other genetic operators like crossover did not incorporate knowledge.

Experiment P_2_ was aimed at showing that: (1) current guideline knowledge incorporation methods for GA are not manageable; and (2) current GAs do not have natural ways of supporting manageable incorporation of guideline knowledge. In P_2_, more knowledge (FCD, harmony rules, personal data, food and Dietary Reference Intakes) was incorporated in genetic operations and population initialisation.

In experiment P_2_, knowledge was formalised into a Prolog knowledge-base which was queried from the Genetic Algorithm. Table IV and Table V show excerpts of the Food Composition Database and Dietary Reference Intakes respectively. The knowledge in the two tables have not yet been formalised into computer interpretable guidelines. Listing I and Listing II show code for nutrition knowledge which was incorporated in experiment P2. Listing I shows the knowledge from the Food Composition Database in Table IV, while Listing II shows a representation of some of the knowledge in Table V

**Table IV:** EXTRACT OF FOOD COMPOSITION DATA BEFORE FORMALISATION (CHITSIKU, 1989)

| Zimbabwe Food list | Energy kCal | Protein | Carbohydrate | Calcium | Iron | Sodium | Zinc | Copper |
| --- | --- | --- | --- | --- | --- | --- | --- | --- |
| Apple | 52 | 0.26 | 13.81 | 6 | 0.12 | 1 | 0.04 | 0.027 |
| Avocado | 167 | 1.96 | 8.64 | 13 | 0.61 | 8 | 0.68 | 0.17 |
| Bananas | 89 | 1.09 | 22.84 | 5 | 0.26 | 1 | 0.15 | 0.078 |
| Baobab | 439 | 5.22 | 1.83 | 35.14 | 29 | 162 | 330 |  |
| Orange | 47 | 0.94 | 11.75 | 40 | 0.1 | 0 | 0.07 | 0.045 |
| Lemon | 29 | 1.1 | 9.32 | 26 | 0.6 | 2 | 0.06 | 0.037 |

**Table V:** DIETARY REFERENCE INTAKES BEFORE FORMALISATION (NATIONAL ACADEMY OF SCIENCES, 2006)

| Life Stage Group | Total Water<sup>a</sup> (L/d) | Carbo-hydrate (g/d) | Total Fiber (g/d) | Fat (g/d) | Linoleic Acid (g/d) | α-Linolenic Acid | Protein (g/d) |
| --- | --- | --- | --- | --- | --- | --- | --- |
| Infants |  |  |  |  |  |  |  |
| 0-6 months | 0.7* | 60* | ND | 31* | 4.4* | 0.5* | 9.1* |
| 7-12 months | 0.8* | 95* | ND | 30* | 4.6* | 0.5* | 11+ |
| Children |  |  |  |  |  |  |  |
| 1-3 years | 1.3* | 130 | 19* | NDc | 7* | 0.7* | 13 |
| 4-8 years | 1.7* | 130 | 25* | ND | 10* | 0.9* | 19 |

| Males |  |  |  |  |  |  |  |
| --- | --- | --- | --- | --- | --- | --- | --- |
| 9-13 years | 2.4* | 130 | 31* | ND | 12* | 1.2* | 34 |
| 14-18 years | 3.3* | 130 | 38* | ND | 16* | 1.6* | 52 |
| 19-30 years | 3.7* | 130 | 38* | ND | 17* | 1.6* | 56 |
| 31-50 years | 3.7* | 130 | 38* | ND | 17* | 1.6* | 56 |
| 51-70 years | 3.7* | 130 | 30* | ND | 14* | 1.6* | 56 |
| >70 years | 3.7* | 130 | 30* | ND | 14* | 1.6* | 56 |

Domain experts recommended use of DRIs. Since DRIs for the United States of America were readily available, they are used in this paper. Meals from P_1_ and P_2_ were compared on quality to demonstrate that GAs do not solve the MPP in their natural form without incorporation of plentiful knowledge. The choice of genetic parameters (chromosome length, population size, crossover and mutation probability) was informed by previous studies which implemented GAs to solve the MPP (see Table VI). 250 generations were used in all experiments, and the crossover probability was 0.9 while the mutation probability was 0.2. In both P_1_ and P_2_, meals were prepared under the assumption that three meals are taken per day by a 35-year old male adult who was physically active.

**Table VI.** EXPERIMENTAL DESIGN PARAMETERS.

| Parameter | Parameter value used in this paper | Parameter values used by other researchers | Parameter | Parameter values used in this paper | Parameter values used by other researchers |
| --- | --- | --- | --- | --- | --- |
| Chromosome Length | 7 | 10 [48]; 5 [49]; 7 [45] | Number of generations | 250 | 6-250 [49]; 40 [22]; Max of 200 [45] |
| Number of meals per day | 3 | 5 [48] | Mutation type | Gaussian | Uniform mutation |
| Population size | 200 | 100 [48]; 40-200 [49] | Crossover type | One point crossover | Linear descending mutation [48]; Arithmetic crossover; Single point crossover [49]; Partially Matched Crossover [29]; Two-step simple crossover [45] |
| Crossover probability | 0.9 | 0.7 [48]; 0.9 [22,49]; | Selection type | Tournament selection | Tournament selection [45]; [48] |
| Mutation probability | 0.2 | 0.1-0.017 [48]; 0.2 [49]; 0.1 [22]; | Population initialisation | Generated randomly | Randomly Generated [49] |
| Number of Iterations | 250 | 35 [48]; 150000 [49]; 10 |  |  |  |

### B Results of Experiments: Management Challenges of Knowledge Incorporation into Solution Models for the MPP

Fig. 3 shows the results from both P_1_ and P_2_ in which 241 meals were produced for each part of the experiment. The mean fitness of P_1_ is 21 while the mean of P_2_ is 17.

#### LISTING I EXTRACT OF FOOD COMPOSITION KNOWLEDGEBASE IN PPROLOG

1. nutrientQuantity(19, avocado, protein, 1.96).
2. nutrientQuantity(12, bananas, protein, 1.09).
3. …
4. nutrientQuantity(18, apple, calcium, 6).
5. nutrientQuantity(19, avocado, calcium, 13).…

**Fig. 3.**
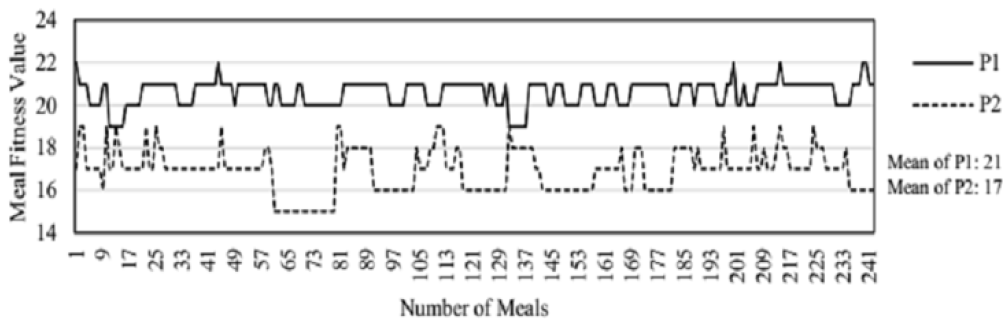
Results of experiments P1 and P2

#### LISTING II DIETARY REFERENCE INTAKES IN PROLOG

1. lifeStageGroup(male).
2. ageGroup(f,31,50).
3. belongsToAgeGroup(X,Y):-ageGroup(X,M,N), Y> = M, Y = <N.
4. nutrient(energy).
5. nutrient(protein).
6. nutrient(carbohydrate).
7. unit(energy, kcal).
8. unit(protein, g).
9. unit(carbohydrate, g).
10. rdi(male, b, protein, 19).
11. rdi(male, f, carbohydrate, 130).
12. #Energy requirements sex(male).
13. age(u,31,35).
14. belongsToage(X,Y):-age(X,M,N),Y> = M,Y = <N.
15. energy(male,u,sedentary,energy,2200).
16. energy(male,u,moderately_active,energy,2400).
17. energy(male,u,active, energy,2800).

#### (1) Results: GAs do not solve the MPP in their natural form without incorporating much knowledge

In P_1_, little knowledge (personal data and FCD) was incorporated in the GA resulting in meals with high fitness values but low levels of harmony as shown in Table 8. Meals from P_1_ have higher fitness values because harmony has been sacrificed. Such meals satisfy most nutrient requirements but are not edible. In P2, more knowledge (personal data, FCD, harmony and Dietary Reference Intakes) was incorporated in the GA resulting in meals with relatively lower fitness values but higher levels of harmony as shown in Table VII. Meals from P1 cannot be classified as either meals for breakfast or lunch or dinner which can be done with meals from P2. In P1, the crossover function does not incorporate knowledge (see Listing II) resulting in crossing over of food items which are not in the same category thereby producing meals with very low levels of harmony. The same conclusion can be reached if the other genetic operations are not knowledge-based.

**Table VII.** SAMPLE MEALS FROM THE EXPERIMENT.

| Experiment | Sample Meals | Fitness value out of 23 | Resulting Meal Harmony | Knowledge Incorporated |
| --- | --- | --- | --- | --- |
| P1 | Powdered milk (136.8g), Okra (230.3g), Cabbage cooked (306.0g), Turnip (200.0g), Chicken with skin (428.0g), Dark bread (162.8g) | 21 | Low | Personal data and some FCD |
|  | Guava (358.0g), Powdered milk (98.9g), Spinach (219.5g), Pumpkin (449.6g), and Cake homemade (184.8g) | 21 | Low |  |
|  | Powdered milk (401.4g), Yogurt (439.7g), Okra (72.5g), Chicken with skin (309.0g), mufushwa (405.4g), Candy (355.2g) | 21 | Low |  |
|  | Guava (212.8g), Powdered milk (68.1g), Cheese Regular (246.2g), Beans or lentils (39.1g), Avocado (251.5g), Lima beans (133.4g) | 20 | Low |  |
| P2 | Spinach (486.5g), Samp (146.5g), Fish (7.0g), Lima beans (249.4g), Mowa (372.0g), Beans or lentils (79.4g) | 19 | High | Personal data, harmony of food items, FCD and Dietary Reference Intakes. |
|  | Spinach (229.7g), Taro (334.0g), Peas, cooked (336.5g), Pumpkin (381.8g), Macaroni, cooked (470.9g), Cake homemade (197.2g) | 19 | High |  |
|  | Cabbage, fresh (300.7g), Pumpkin (223.1g), Birds (22.3g), Sadza from sorghum (351.0g), skimmed milk (152.9g), Canned tuna (43.8g) | 19 | High |  |
|  | Carrot, fresh (263.7g), Okra (364.9g), Samp (90.4g), Avocado (419.2g), Fish (303.1g), Lima beans (450.4g) | 19 | High |  |

**Table VIII.** GENETIC ALGORITHM MODIFICATION WHEN KNOWLEDGE IS INCORPORATED.

| Genetic Operation | Experiment P<sub>2</sub> |  |
| --- | --- | --- |
|  | GA Code Change | Genetic Algorithm Modification |
| Crossover | ✓ | Function modified to crossover only food items in the same category |
| Mutation | ✓ | Function modified to ensure food item weights did not exceed limits |
| Fitness function | ✓ | Function modified to incorporate FCD and NRVs |
| Selection | ✓ | Only food items in the same category were selected |

In summary, GAs do not solve the MPP in their natural form without incorporating much knowledge. This finding confirms what is in found in literature since there are some works [46], [22], and [48] which incorporated domain knowledge in GAs for the MPP even though the knowledge was hard to manage. However, there are very few works in literature which modelled the MPP using GA but without incorporating much domain knowledge such as [43]. As can be seen these works are now aged and the current trend is to incorporate domain knowledge in GA in order to solve the MPP.

#### (2) P_2_ Results: The difficulty of Managing Incorporated Knowledge

##### GAs do not have natural ways of supporting incorporation of knowledge

Knowledge can be incorporated into GAs by either tightly integrating into the genetic operators or storing in a knowledge-base to be *consulted* by the operators. Both approaches will require provision for query and operator support for managing the knowledge, which is widely lacking in the research literature. For example, in Experiment P2, the crossover function had to be modified to incorporate knowledge about food item categories otherwise unpalatable meals were going to be produced. Table VIII shows how Genetic operators were also modified to incorporate domain knowledge. The same table also shows the Genetic operations and associated changes which were undertaken to incorporate knowledge. In VIII it is shown that the GA code had to be changed in order to incorporate knowledge in the algorithm. A tick (⌞) in the table means change that the GA code had to be changed to implement the function. This result is supported by [47] who had to devise a way of replacing harmony rules by implementing them as a plug-in. Even though this was good idea, it is far from being adequate for supporting querying and manipulation of the incorporated knowledge.

##### Knowledge incorporated into GA-based models for the MPP using existing approaches is hard to manage

The previous result has shown that GAs indirectly support knowledge incorporation. However, the knowledge incorporated using the existing approach is hard to manage if there is *no support* for query and manipulation operators on the incorporated knowledge. In P2, it is not easy to manage the incorporated knowledge because the model does not support management operations such as *create, update, delete, customise, replace* and *query*. Table IX presents the manipulation and query operations on incorporated knowledge indicating whether or not changes must be performed on the Genetic Algorithm code and the incorporated knowledge when the operators are applied. A tick (✓) in Table IX means change is necessary for the management operation to be applied, while a blank cell means no change is required. A question mark (?) means the operation cannot the applied to the GA or knowledge. Table IX also shows that if management operations are to be applied to P2 then both the knowledge and model should change. This *model-knowledge* dependency makes it hard to manage knowledge incorporated into GAs for the MPP using the existing approaches.

**Table IX.** CHANGES TO BE EFFECTED WHEN MANAGEMENT OPERATIONS ARE APPLIED.

|  | Management Operation | Experiment P<sub>2</sub> |  |
| --- | --- | --- | --- |
|  |  | GA Code Change | Knowledge-Base Change |
| Standard CRUD Operations on Knowledge | Create | ✓ | ✓ |
|  | Retrieve/Query |  |  |
|  | Update/Change | ✓ | ✓ |
|  | Delete/Remove | ✓ | ✓ |
| Advanced knowledge manipulation operations | Customise | ✓ | ✓ |
|  | Replace | ✓ | ✓ |
|  | Share | ? | ? |

## VII TOWARDS SUPPORTING THE MANAGEMENT OF INCORPORATED KNOWLEDGE

The work of the authors aim to create a foundation for an approach to manageable incorporation of knowledge into solution models incorporated into technologies that may be transferred to developing regions of the world. The main challenge is to achieve knowledge and model independence.

Thus, if the knowledge changes, the code or model does not have to be modified. Several works in literature like [22] and [48] report attempts to incorporate knowledge into the GA but the knowledge was not *highly targeted knowledge* (i.e., not problem-specific) and did not define the problem of supporting the customisation of the incorporated knowledge. In the absence of such support and the existence of the need to manipulate the knowledge, the resulting technology cannot be customised. *A new approach is required in which there is comprehensive support for the management of the incorporated knowledge without affecting the solution models to facilitate appropriate technology transfers*.

How manageable incorporation of *highly targeted* food and nutrition therapy guideline knowledge into solution models for the MPP can be achieved remains a challenge that is worthy of further research attention. For knowledge incorporated into a solution model for the MPP to be useful and manageable, the knowledge must be *highly targeted* and be able to be *queried and manipulated to the lowest granularity* possible.

Most solution models require very tight coupling between the model and the incorporated knowledge elements in order to produce *realistic solutions*. Therefore, *on-going work* of the authors will explore an approach based on the combination of *knowledge and model-based methods with formal process and model specifications*, which is expected to allow both the solution model and the incorporated knowledge to be formally *specified, executed, queried* and *manipulated*. Even when the solution model and the incorporated knowledge are tightly integrated, both are expected to be subjected to *query, execution* and *manipulation* operations leading to technologies that are customisable to suit local circumstances.

## VIII SUMMARY

This paper investigated the challenge of managing knowledge incorporated into computational models. This was done in the context of manageable incorporation of food and nutrition therapy guideline (FNTG) knowledge into genetic algorithms (GA) for the meal planning problem (MPP). A framework for manageable incorporation of knowledge into the solution model based on the GA for the MPP was developed. Experiments were undertaken to investigate how manageable the knowledge incorporated in the GA for the MPP is. Results revealed that the incorporated knowledge the GA solution model is difficult to manage. This conclusion could be extended to solution models other than the models used in this paper and to other problem domains than the MPP.

New engineering methods that support operations to execute, query and manipulate knowledge incorporated into solution models would be beneficial in the development of customisable technologies that would be transferrable across regions of the world.

Future work will entail: (1) Developing a new approach to manageable incorporation of formalised highly targeted knowledge into solution models for the MPP; (2) Developing strategies and better engineered solutions for real uses of the MPP solution model in the context of HIV/AIDS nutrition therapy; and, (3) Application of the model in the mobile web-based context of a developing country to facilitate effective manageable knowledge and technology transfer from experts to people living with HIV/AIDS.

## Data Availability

not applicable

## ACKNOWLEDGEMENTS

The authors would like to acknowledge: *University of Zimbabwe* (UZ) for supporting the work of NZ, *Massey University* for supporting JST work and funding KD’s sabbatical visit to UZ and *Queen Mary University of London* (QMUL) during Jan – Feb, 2019. SM, EK and NF acknowledge support from QMUL and the *EPSRC* under EP/P009964/1: PAMBAYESIAN: *Patient Managed Decision-Support Using Bayes Networks*.

## Notes

### Competing Interest Statement

The authors have declared no competing interest.

### Clinical Trial

This work is neither a clinical trial nor prospective study but purely Health Informatics research project. There was not need to register.

